# Early estimates of SARS-CoV-2 B.1.1.7 variant emergence in a university setting

**DOI:** 10.1101/2021.03.05.21252541

**Authors:** Kaitlyn E. Johnson, Spencer Woody, Michael Lachmann, Spencer J. Fox, Jessica Klima, Terrance S. Hines, Lauren Ancel Meyers

## Abstract

Recent identification of the highly transmissible novel SARS-CoV-2 variant in the United Kingdom (B.1.1.7) has raised concerns for renewed pandemic surges worldwide ^1,2^. B.1.1.7 was first identified in the US on December 29, 2020 and may become dominant by March 2021 ^3^. However, the regional prevalence of B.1.1.7 is largely unknown because of limited molecular surveillance for SARS-CoV-2 ^4^. Quantitative PCR data from a surveillance testing program on a large university campus with roughly 30,000 students provides local situational awareness at a pivotal moment in the COVID-19 pandemic.

## Methods

We estimate the prevalence of B.1.1.7 based on daily positive SARS-CoV-2 PCR tests and the number of S gene target failures (SGTF) out of 17,003 student tests reported by the University of Texas’ Proactive Community Testing (PCT) between January 16 and February 12, 2021. Mutations in the spike protein of the B.1.1.7 variant result in a failure to detect the S gene probe in a standard SARS-CoV-2 RT-qPCR test. In January 2021, the proportion of B.1.1.7 variants among sequenced SGTF samples was estimated at ∼90% in the U.S., with substantial regional variance ^4^. Assuming this proportion, we use a Bayesian model to estimate the growth rate of B.1.1.7 prevalence among all SARS-CoV-2 infections at UT and then apply a compartmental SEIR model of SARS-CoV-2 transmission to project the impact of B.1.1.7 on future COVID-19 burden.

## Results

We estimate that the relative frequency of B.1.1.7 is growing logistically at a daily rate of 0.077 [95% CI: 0.017–0.140] which corresponds to an early doubling time of 9.0 days [95% CI: 5.0–41.0 days]. This is consistent with growth rates estimated in the UK (0.072) ^5,6^, Florida (0.076), and California (0.057) ^1,4^.

The estimated proportion of SARS-CoV-2 infections caused by the B.1.1.7 variant in the UT community increased from 2.7% [95% CI: 0.7–7.6%] on January 16 to 17.9% [95% CI: 9.6–29.4] on February 12. B.1.1.7 is expected to comprise a majority of cases at UT by March 5 [95%CI: February 20–March 28]. Assuming B.1.1.7 is 1.56 times as transmissible as the wildtype ^1^, we project that its emergence will lead to 77% [95%CI: 39–140%] more cumulative infections during the spring semester than in the absence of the variant.

## Discussion

Surveillance testing conducted by a large public university located in a metropolitan area with a population over two million indicates that B.1.1.7 is rapidly becoming the dominant variant and has the potential to drive a major surge in infections during the spring 2021 within a community not likely to be widely vaccinated before summer 2021. Our methodology allows early detection of B.1.1.7 emergence from widely available quantitative PCR data, when sequence confirmation is not available or delayed, while quantifying uncertainty in the variant growth rate and fraction of SGTF samples that are positive for B.1.1.7. Notably, between January 16–February 5, four SGTF SARS-CoV-2 specimens were sequenced by UT and all were confirmed positive for B.1.1.7, corroborating our reliance on SGTF data.

While our estimates are derived from limited surveillance data, they are consistent with trends observed in the UK and other US states and reinforce the urgent need for expanded molecular surveillance capacity and continued COVID-19 mitigation efforts in universities and surrounding communities. Until sequencing is more widely available, quantitative PCR data from large-scale testing efforts can provide sentinel warning of B.1.1.7 emergence in cities throughout the US.

For this study, we assume that daily SARS-CoV-2 specimens are from a representative sample of the UT community and that B.1.1.7 constitutes a fixed proportion of SGTF specimens over time. We discuss possible biases resulting from these assumptions in the supplement.

**Figure 1.**
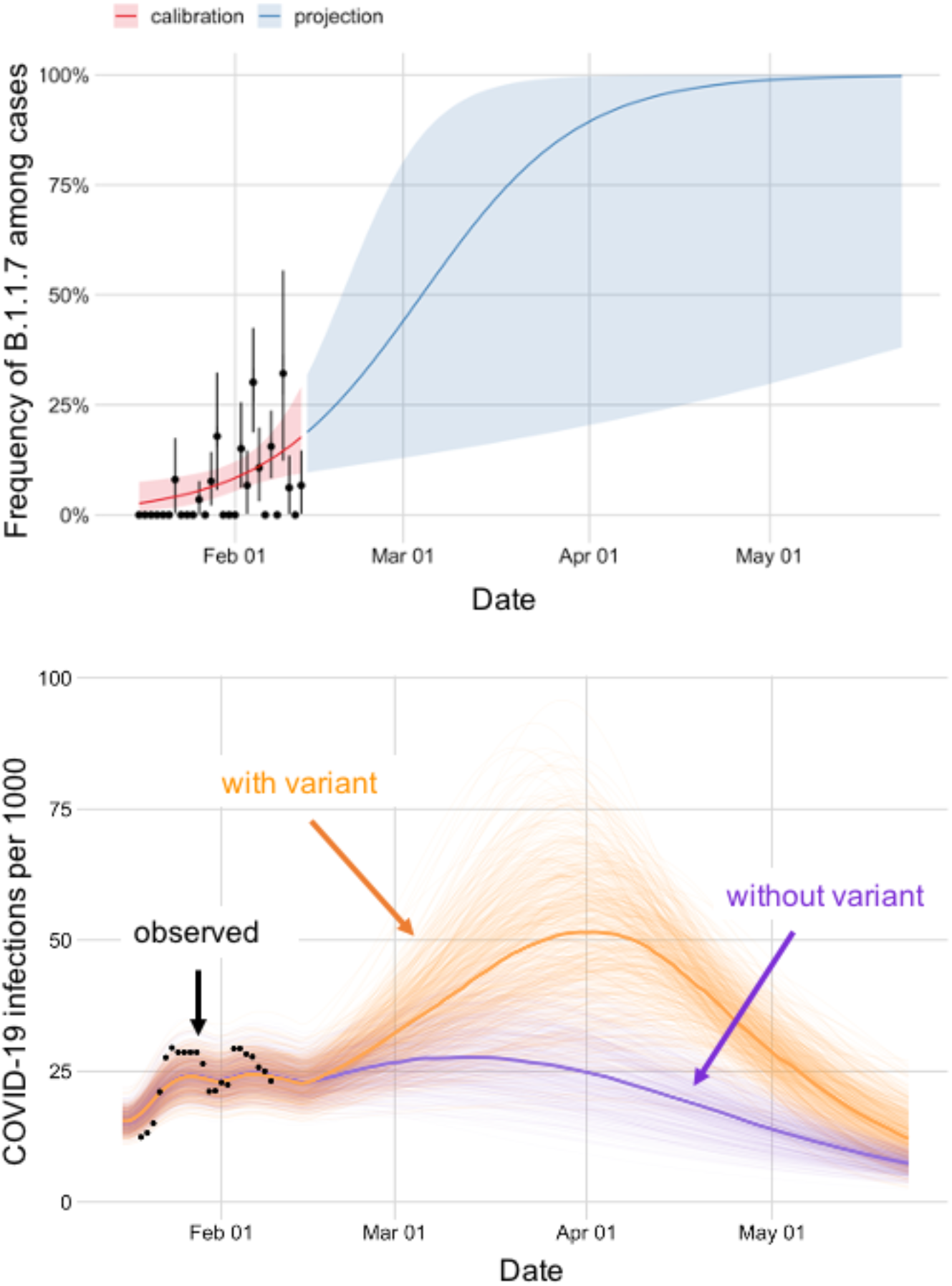
Estimated frequency of the B.1.1.7 variant among positive COVID-19 cases at the University of Texas and its projected impact on COVID-19 prevalence, January 16 - May 23, 2021. A. Based on the number of samples with SGTF among SARS-CoV-2 positive samples reported by UT Proactive Community Testing (PCT), we estimate the frequency of the B.1.1.7 variant (black points). Vertical error bars represent standard errors. The calibrated logistic growth model (red) and projections from the fitted model (blue) indicate rapid spread of the B.1.1.7 variant relative to the previously circulating (wildtype) virus. Shaded bands indicate 95% credible intervals, which reflect uncertainty in the percent of cases that are S gene dropouts, the percent of S gene dropouts that are B.1.1.7, and the fitted model parameters. B. Projected COVID-19 cases at UT throughout the spring semester of 2021. Orange and purple indicate projections with and without the variant, respectively, and the black dots indicate the seven-day average reported positive cases detected through UT Proactive Community Testing (PCT) per 1000. The projections assume *R*_*t*_ = 1.17 [95%CI: 0.97-1.39] as of February 12, based on the *R*_*t*_ as of February 5. Light lines display 500 simulations; bold lines indicate the median projected value on each day. Additional scenario projections can be found in the Supplement.

## Data Availability

Test positivity data used in this manuscript is available publicly at the UT Austin COVID-19 Dashboard.
Use of deidentified quantitative PCR data was approved by the IRB at UT Austin.

https://coronavirus.utexas.edu/ut-austin-covid-19-dashboard

