## Supplement for "Early estimates of SARS-CoV-2 B.1.1.7 variant emergence in a university setting"

#### Supplementary table and figure

**Table S1. Weekly SGTF and total positive samples reported by UT PCT and estimated percent of COVID-19 cases that are infected by the B.1.1.7 variant in the UT community.** Estimates are given as posterior medians and 95% credible intervals for the Friday of the week indicated. Bold rows correspond to future projections based on the observed trend through February 12, 2021.

| | Samples with SGTF | Total COVID-19 positive samples | Estimated percent of cases infected by B.1.1.7 variant* |
|---|---|---|---|
| **Estimates** | | | |
| Jan. 16 - 22 | 1 | 49 | 4.2% [1.7-8.6%] |
| Jan. 23 - 29 | 5 | 93 | 6.9% [4.0-10.6%] |
| Jan. 30 - Feb. 5 | 15 | 75 | 11.2% [7.5-15.9%] |
| Feb. 6-12 | 10 | 79 | 17.9% [9.6-29.4%] |
| **Projections** | | | |
| **Feb. 13 - 19** | NA | NA | **27.3% [11.2-51.2%]** |
| **Feb. 20 - 26** | NA | NA | **39.1% [12.7-73.3%]** |
| **Feb. 27 - Mar. 5** | NA | NA | **52.3% [14.2-87.9%]** |
| **Mar. 6 - Mar. 12** | NA | NA | **65.2% [15.9-95.1%]** |
| **Mar. 13 - Mar. 19** | NA | NA | **76.3 [17.6-98.1%]** |
| **Mar. 20 - Mar. 26** | NA | NA | **84.6% [19.5-99.3%]** |

*Estimated for Friday of the specified week.

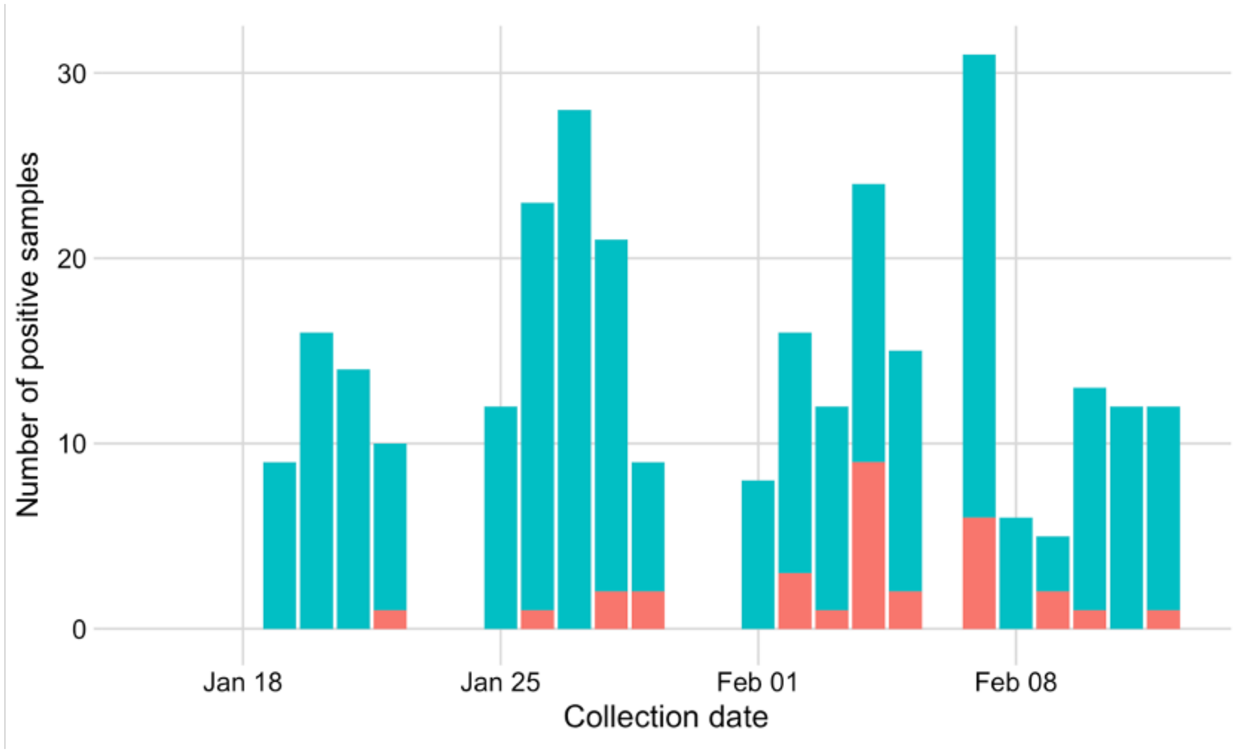


**Figure S1. Daily numbers of positive SARS-CoV-2 samples** with Ct<28 reported by UT PCT from January 16 to February 12, 2021, stratified by non-SGTF (blue) and SGTF (red).

#### RT-qPCR data from university testing program

We analyzed de-identified lab results from the Proactive Community Testing (PCT) program at the University of Texas at Austin [1]. Saliva specimens were collected from individuals at the university presenting for voluntary asymptomatic testing [2]. PCT test results are based on the Thermo Fisher TaqPath$^{TM}$ COVID-19 Combo Kit, which targets three SARS-CoV-2 viral regions (N gene, S gene, and ORF1ab). Since samples are deidentified prior to analysis and some individuals may test more than once, there may be duplicate individuals in the data that could bias our estimates. Test results from positive cases, together with sample collection date and reverse transcriptase quantitative polymerase chain reaction (RT-qPCR) cycle threshold (Ct) values for all gene targets were used to build the dataset. Ct refers to the number of cycles needed to amplify viral RNA to reach a detectable level. Ct values are inversely related to the amount of virus in a specimen.

Specimens are considered SARS-CoV-2 positive when at least two of the three target genes (N, Orf1ab, and S) are detected at a Ct value below 37. Following approaches from prior studies [3,4], we filtered our dataset for positive samples with strong amplification of the N gene (Ct < 28) to increase the sensitivity and specificity of SGTF detection.

S gene target failures occur when RT-qPCR fails to detect the virus' S gene and are caused by mutations in the gene. Deletions in the amino acids H69 and V70 in the B.1.1.7 variant result in an SGTF. Samples were considered to be SGTF samples if they were positive for both N and Orf1ab, and negative for S. While SGTF can occur due to other mutations, the presence of the SGTF is one of several mutations that distinguish the B.1.1.7 variant from other strains [5,6]. All of the SGTF specimens are sent for additional confirmation via sequencing through the university, but sequencing results are often delayed by one to two weeks. Given the need for rapid estimation of variant prevalence, we did analyze not the limited sequencing data that were available at the time of this study. Specifically, only four of the 31 SGTF identified as of February 12th had been sequenced.

Our analysis of B.1.1.7 variant prevalence focuses on the number of positive samples with SGTF out of the total number of high quality (Ct<28) positive SARS-CoV-2 samples collected through PCT. In the US, approximately 70-90% of SGTF samples were confirmed as variants in mid-January 2021 [3]. We use this national proportion in estimating the prevalence of B.1.1.7 based on the SGTF data. Our method includes a prior (beta) distribution governing the proportion of SGTFs that are B.1.1.7 that can be easily updated as new estimates become available, as described in the next section.

#### Projecting B.1.1.7 frequency using a logistic growth model

To estimate the relative frequency and growth of the B.1.1.7 variant, we implement a Bayesian logistic growth model using default priors in the rstanarm package in the R programming language [7]. To start, let $S_t$ be the number of positive case samples with SGTF and low Ct, $B_t$ be the (unknown) number of B.1.1.7 cases at time $t$, and $N_t$ be the total number of positive case samples.

The goal is to estimate the prevalence of B.1.1.7, that is, the percentage of COVID+ cases which contain the variant at time $t$, which we denote by $p_{t,NB}$. Ideally, we would like to sequence the positive cases to detect B.1.1.7, in which case we would assume each COVID+ sample has a $p_{t,NB}$ probability of being B.1.1.7+, so then the number of B.1.1.7+ samples can be described by a binomial distribution

$$B_t \sim \text{Binomial}(N_t, p_{t,NB})$$

Previously, the growth in prevalence of the B.1.1.7 in other countries has closely followed a logistic curve [3], so then the prevalence may be described to evolve over time given by the logistic equation

$$\log \frac{p_{t,NB}}{1 - p_{t,NB}} = \beta_0 + \beta_1 t$$

Here, $\beta_1$ is the growth rate and $\beta_0$ is an intercept term. These coefficients can be estimated using existing regression software implementations. However, the main problem is that we do not know the true number of B.1.1.7 samples. Instead we will impute this number using the number of SGTF samples, and the proportion of SGTF samples $p_{SB}$. This proportion is also not definitively known, so we integrate over uncertainty in estimating the prevalence of the variant. We describe uncertainty in the fraction of B.1.1.7 samples to total SGTF samples by a beta distribution

$$p_{SB} \sim \text{Beta}(40, 10)$$

The parameters of this beta distribution were selected so that the 95% central credible interval is approximately (0.7, 0.9), consistent with the range of findings reported in [3] of percent of B.1.1.7 among S gene dropouts during mid-January 2021 in a number of U.S. states. Estimates for the state of Texas were not available at the time of this study, given the limited capacity for molecular surveillance [3].

We implement the logistic regression binomial sampling model for $B_t$ as described above, integrating over the uncertainty in $p_{SB}$ via Monte Carlo sampling. One Monte Carlo draw of this model works as follows

1. Draw from the beta distribution described above for $p_{SB}$ the fraction of S gene dropouts that are positive for B.1.1.7
2. Impute B.1.1.7 cases by multiplying S gene dropout cases by the draw from the beta distribution
3. Estimate the logistic growth model using this set of imputed B.1.1.7 case numbers,
4. Finally, project future B.1.1.7 prevalence using the fitted model

We combine all draws for projected B.1.1.7 prevalence to integrate over uncertainty in the fraction of B.1.1.7 to S gene dropout samples.

### Projections of COVID-19 spread at UT, spring 2021

We projected the spread of the B.1.1.7 and original (*wildtype*) variants at UT throughout the spring semester of 2021 using a two-variant epidemiological model (Figure S2), described by the equations below. The model assumes that the wildtype and variant strains infect a shared pool of susceptibles, all of whom are assumed to be well-mixed within the UT student community. The model assumes that all individuals infected with either the wildtype or variant strain are fully immune from infection by either strain after recovery. Individuals transition between the states: susceptible (S), exposed (E), infected (I), and recovered (R). The V and W subscripts in the E and I compartments refer to whether the individual is infected with wildtype SARS-CoV-2 (W) or variant SARS-CoV-2 (V), in this case the B.1.1.7 variant. The symbols S, $E_W$, $E_V$, $I_W$, $I_V$, and R denote the number of people in that state. The model equations are given by:

$$\frac{dS}{dt} = -\beta(t)(I_W + p_v I_V)\frac{S}{N}$$
$$\frac{dE_W}{dt} = \beta(t) I_W \frac{S}{N} - \gamma E_W$$
$$\frac{dI_W}{dt} = \gamma E_W - \delta I_W$$
$$\frac{dE_V}{dt} = \beta(t) p_V I_V \frac{S}{N} - \gamma E_V$$
$$\frac{dI_V}{dt} = \gamma E_V - \delta I_V$$
$$\frac{dR}{dt} = \delta I_W + \delta I_V$$

where $\beta$ *(t)* is the baseline transmission rate, $p_v$ is the relative transmissibility of the variant, $\gamma$ is the exposed rate, and $\delta$ is the recovery rate. The initial conditions are given in Table S2, the model parameters are given in Table S3, and the scenario parameters are given in Table S4.

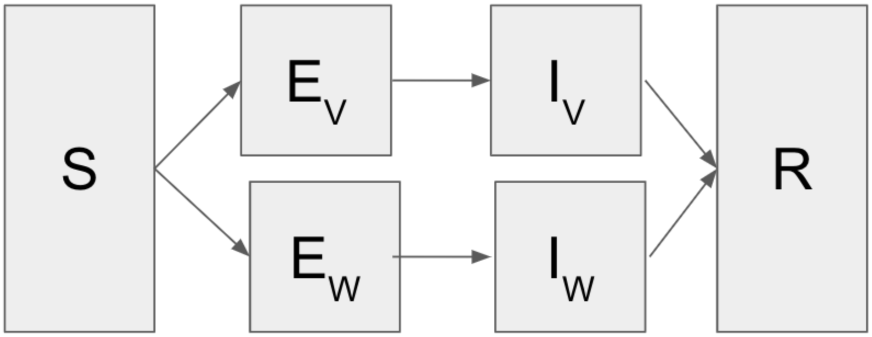


**Figure S2. Diagram of the two-strain COVID-19 transmission model.** Upon exposure to either strain, susceptible individuals (S) progress to either exposed to the wildtype ($E_W$) or exposed to the variant ($E_V$), from which they move to either infected by the wildtype ($I_W$) or infected by the variant ($I_V$) respectively. All infected individuals progress to the recovered state where they remain protected from future infection (R).

**Table S2. Initial conditions for COVID-19 transmission simulations.**

| **Variable** | **Value** |
|---|---|
| Initial day of simulation | 1/16/2021 |
| Day of variant introduction | 2/12/2021 |
| Prevalence of B.1.1.7 variant on day of introduction | 17.9% [95%CI: 9.6-29.4%] of cases are infected by B.1.1.7 |
| Initial proportion infected | 1.56% [95% CI: 1.25- 1.92%] are infected |
| Initial proportion immune (percent of students previously infected, as estimated from fall UT testing data) | 14.7% [95% CI: 10.7-20.9%] of UT students are immunized from prior infection [1,8] |

**Table S3. Model parameters**

| Parameters | Value | Source |
|---|---|---|
| $N$: Number of UT students in Austin | 30,000 | [9] |
| $\gamma$: transition rate from exposed to infectious | 1/3 | [10] |
| $\delta$: recovery rate | 1/7 | [11] |
| $p_v$: relative transmissibility of the variant | 1.56 (1.50-1.74) | [12] |
| $R_t$: reproduction number | Slower scenario: 0.99 [95%CI: 0.81-1.19] Faster scenario: 1.17 [95%CI: 0.97-1.39] | Estimated using EpiEstim [8] from UT PCT data [1] |
| $\beta$ (t): transmission rate | Slower scenario: 0.19 [95% CI: 0.16, 0.24] Faster scenario: 0.23 [95% CI: 0.19, 0.28] | Calculated from $R_t$ |

**Table S4. Scenario parameters**

| **Transmission scenarios after February 12** | |
|---|---|
| **All eight combinations of these three factors** | The wildtype spreads either slower ($R_t$ = 0.99 [95%CI: 0.81-1.19]) or faster ($R_t$ = 1.17 [95%CI: 0.97-1.39]) |
| | The B.1.1.7 variant either does or does not spread alongside the wildtype |
| | Spring break either does or does not increase transmission by 100% for the four days following the break (March 20 – 23) |

#### Initial conditions

For each simulation, we sample from the distribution of proportion immune (previously infected) and the distribution of the proportion arriving to campus infected. The initial number of infected students is estimated using the observed number of positives out of the total number of tests administered to students over the first week of testing (January 16th - January 22nd). If we assume each individual has a probability $p_{t=0,inf}$ of being infected, the number of observed positives, $N_+$, out of the number of tests administered, $N_{tests}$, can be described by a binomial distribution:

$$N_+ \sim Binomial(N_{tests}, p_{t=0,inf})$$

If we assume a flat beta prior on the probability of being infected at t=0 of $p_{t=0,inf} \sim Beta(1,1)$, we can write the posterior probability of an student being infected as:

$$p_{t=0,inf} \sim Beta(1 + N_+, 1 + N_{tests} - N_+)$$

Using the number of positives and the total number of tests from the first week, we draw from the posterior distribution of the probability of a student being infected, then impute the initial number infected by multiplying by the total number of students at UT.

The assumed distribution of proportion immune is based on (i) estimates for the number of cases in August based on reported COVID-19 incidence in the residential counties of 30,000 returning students [13] and (ii) $R_t$ values from August 20th to December 19th, 2020 estimated from all student positive cases using the EpiEstim package [8]. Using a single-strain version of the SEIR model and workflow presented below, the daily transmission rate $\beta$ was calculated directly from $R_t$. The total number of cumulative infections for each simulation was used to estimate the initial number of individuals in the recovered compartment at the start of the spring semester. We assumed 2% of the student body was recovered and immune prior to the fall semester and did not account for additional infections that occurred during the winter break. If prior immunity was much higher than our estimates, then our projections would overestimate prevalence throughout the spring. If immunity is incomplete, allowing for some level of reinfection by the wildtype or B.1.1.7 variant, then our projections may underestimate the potential surge.

#### SARS-CoV-2 Transmission Rate

We estimate the SARS-CoV-2 transmission rate among UT students between January 16 and February 12, 2021 based on publicly available daily numbers of positive tests and total tests administered by PCT [1]. For this analysis, we did not exclude positive cases with high Ct values. To correct for fluctuations in testing levels, we calculate the “test-level corrected” case count as the total number of positive cases divided by the total number of tests administered, multiplied by the average number of tests per day administered between January 16 and February 12.

The daily reproduction number ($R_t$) prior to February 12 is sampled directly from estimates of the distribution of $R_t$ from the test-level corrected case count, using the EpiEstim package [8]. $R_t$ is calculated over a 7 day window, resulting in $n$-7 $R_t$ estimates for $n$ days of case data. We assumed $R_t$ remained the same value as was observed on February 5th to fill in the transmission rate from February 5th-12th. After February 12th, we projected 8 scenarios which assume a fixed transmission rate (except for the spring break surge) sampled from a distribution of $R_t$ corresponding to either the slower transmission or faster transmission scenario. These were chosen from recently observed $R_t$ values. The transmission rate ($\beta$) corresponding to the specified $R_t$ is then given by:

$$\beta(t) = R_t \delta \frac{N}{S(t)}$$

In each simulation, $\beta$ (t) is held constant after February 12th, allowing $R_t$ to decline as the pool of susceptibles are depleted.

#### Projected infections under each scenario

The projections suggest that the rapid emergence of the B.1.1.7 variant in January and February would lead to much higher COVID-19 prevalence in the UT community throughout the spring semester, even if the overall transmission is reduced through mitigation (Figure S2). In the worst-case scenario (high transmission with a spring break surge), we would expect the pandemic wave to peak towards the end of March, with the B.1.1.7 variant nearly doubling the number of infections at the peak (72 [95%PI: 35-112] cases per 1000 with the variant versus 36 [95%PI: 22-60] cases per 1000). We would also expect the B.1.1.7 variant to nearly significantly increase the total number of students infected between January 16 and May 23 under this worst-case scenario from 13,018 [95%PI: 8,923, 17,153] to 19,502 [95%PI: 14,462-22,355].

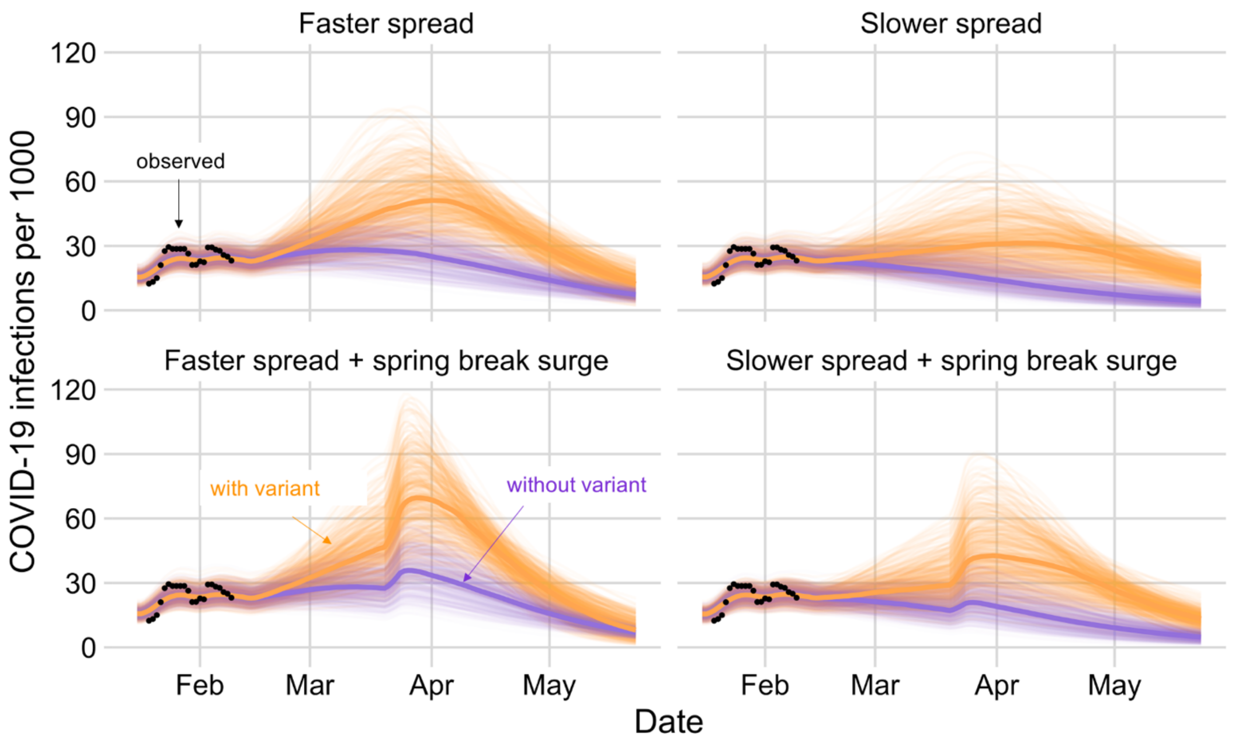


**Figure S3. Projected COVID-19 cases at UT throughout the spring semester of 2021 under eight transmission and variant scenarios.** In all graphs, orange and blue indicate projections with and without the variant, respectively, and the black dots indicate observed cases detected through UT Proactive Community Testing (PCT) per 1000 (seven-day average). The left column of graphs show projections under the faster transmission scenarios ($R_t$ =1.17 [95%CI: 0.97-1.39]) with (top) and without (bottom) a post spring break surge; the right graphs show the corresponding projections under the slower transmission scenario ($R_t$ =0.99 [95%CI:0.81-1.19]). For the spring break surge, we assume the transmission rate doubles from March 20 to 23. For each scenario, we display 500 simulations, with the bold line indicating the median projected value on each day.

#### Additional Limitations

In addition to the limitations listed in our letter, we make the following assumptions. First, we assume that PCT testing represents a random sample of the entire student population. However, testing is voluntary. If students are more likely to test after known exposures, then we may overestimate initial prevalence. Alternatively, if students engaging in riskier behaviors are less likely to seek testing, then we may underestimate prevalence.

Second, we assume that SGTF prevalence among positive PCT specimens is representative of SGTF prevalence in the UT Community as whole. However, the location of PCT testing varies each day and is sometimes targeted towards certain populations, and therefore cases tend to cluster geographically by day [2]. These two factors might increase the chance of detecting a cluster of related B.1.1.7 cases that are not indicative of the overall prevalence of the variant in the UT community. This could lead us to overestimate both its local prevalence and growth rate. However, we note that it is unlikely that B.1.1.7 cases are being systematically selected for testing within the data up to this point. All tests collected prior to February 5, 2021 at UT occurred before sequencing confirmation of the presence of B.1.1.7 on campus, and no effort was made to perform more aggressive contact-tracing of these individuals prior to this date.

Third, we note that our indirect estimates of immunity and transmission rates within the UT community are based on limited data from the fall semester of 2020, and thus are highly uncertain.

Fourth, we assume that the B.1.1.7 variant will have a transmission advantage over the wildtype variant based on estimates from the UK. The transmission rate of B.1.1.7 in Austin may differ from these estimates, as it will depend on the extent of individual and community efforts to slow transmission as well as the levels of infection-acquired and immune-acquired immunity, which may differ from conditions in the UK during November and December 2020 period.

Finally, we make the simplifying assumption that infection by either variant renders an individual immune to reinfection by either variant, despite a number of reports of COVID-19 reinfections [14]. While reinfection may become more likely as the virus continues to evolve, scientists believe that past infections provide a reasonable degree (but not full) immunity and that reinfections are not a primary driver of B.1.1.7 transmission [12].

### Pairwise analysis of Ct values among non-SGTF and SGTF positive SARS-CoV-2 samples

In order to test the hypothesis that the B.1.1.7 variant's increase in transmissibility may be due to higher viral load of individuals infected with the variant, we examined the distribution of Ct values amongst non-SGTF and SGTF samples from all SARS-CoV-2 positive tests. The Ct value, or cycle threshold, refers to the number of cycles needed to amplify the 3 target genes (N, S, and ORf1ab) of SARS-COV-2 to detectable levels. A higher Ct value indicates a lower viral load. In order to ensure that the SGTFs analyzed were due to deletions in viral RNA rather than too little viral presence, we previously restricted our analysis of B.1.1.7 prevalence to samples with mean Ct values over all 3 genes or the remaining genes if S gene was not detected (dashed line, Figure S4).

However, in order to investigate whether SGTF positives have systematically higher viral loads (lower Ct values) we looked at the Ct value of all available SARS-COV-2 positive samples collected from January 16, 2021 to February 12th, 2021 (Figure S4). In total, there were 39 SGTF positives and 364 non-SGTF positives. Of the 39 SGTF positives, 31 (79.5%) of them had Ct values less than 28. Of the 364 non-SGTF positives, 265 (72.8%) of them had Ct values less than 28.

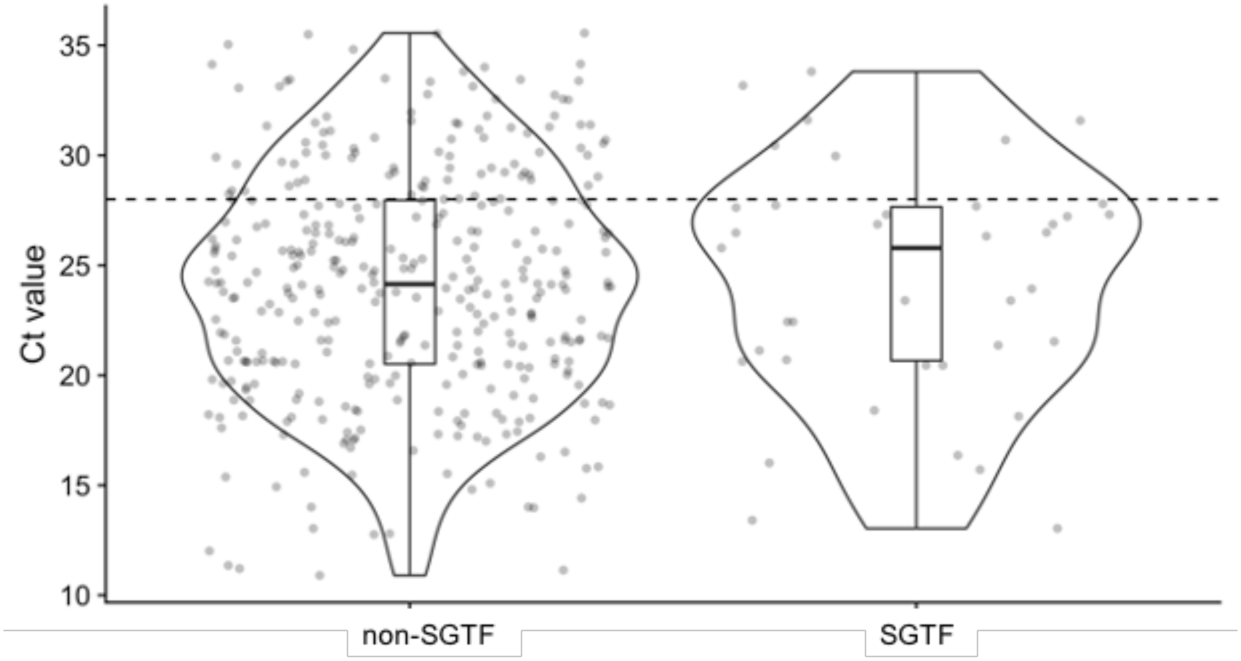


**Figure S4. Ct values of non-SGTF vs. SGTF positive SARS-COV-2 samples collected from PCT from January 16, 2021 to February 12, 2021.** Dashed line at Ct=28 indicates the threshold value below which positive SARS-CoV-2 samples were included in the estimates of B.1.1.7 prevalence. Box plots indicate the median and quartiles over all Ct values for each group. Dots indicate individual sample Ct values. Violin plots show density of samples at each Ct value.

Analyzing all of the SGTF and non-SGTF positives, we find that the median Ct value of non-SGTF positives is 24.1 (95%CI: 14.0-33.5), while the median Ct value of the SGTF positives is 25.8 (95%CI: 13.4- 33.2). To test for significant differences in these two distributions, we ran a two-sample Kolmogorov-Smirnov test on the Ct values of the non-SGTF and SGTF samples, using the ks.test function in R, giving $p = 0.4125$. We thus see no statistically significant difference between the two distributions. We find no evidence within this small data set that SGTF positive individuals have higher viral loads than individuals with non-SGTF positives.
